# E-cigarette use and combustible tobacco cigarette smoking uptake among non-smokers, including relapse in former smokers: umbrella review, systematic review and meta-analysis

**DOI:** 10.1101/2020.09.16.20195438

**Authors:** Olivia Nina Baenziger, Laura Ford, Amelia Yazidjoglou, Grace Joshy, Emily Banks

**Author notes:** **Corresponding author**: Emily Banks.

## Abstract

Combustible tobacco smoking is a leading cause of death and disability worldwide. E-cigarettes are promoted for smoking cessation, but evidence on how their use relates to smoking uptake is limited. We searched PubMed, Scopus, Web of Science, PsychINFO (Ovid), Medline (Ovid) and Wiley Cochrane Library in April 2020. Studies of non-smokers - never, not current, and former smokers - with a baseline measure of e-cigarette use and an outcome measure of combustible smoking uptake were included. Of 6,225 studies identified, 3 systematic reviews (incorporating 13 primary research studies) and 12 additional studies were included in umbrella and top-up systematic reviews, respectively. All 25 studies found increased risk of smoking uptake with e-cigarette exposure, although magnitude varied substantially. Using a random-effects model, comparing e-cigarette users versus non-e-cigarette users, among never-smokers at baseline the odds ratio (OR) for smoking initiation was 3.25 (95%CI 2.61-4.05, I^2^ 85.7%) and among non-smokers at baseline the OR for current smoking was 2.87 (95%CI 1.97-4.19, I^2^ 90.1%). Among former smokers, smoking relapse was higher in e-cigarette users versus non-users (OR=2.40, 95% CI 1.50-3.83, I^2^ 12.3%). Across multiple settings, non-smokers who use e-cigarettes are consistently more likely than non-e-cigarettes users to initiate combustible cigarette smoking and become current smokers; risk magnitude varied, with an average of around three times the odds. Former smokers using e-cigarettes have over twice the odds of relapse as non-e-cigarettes users. This study is the first to our knowledge to review and pool data on the latter topic.

## Introduction

Globally, combustible tobacco smoking results in over 8 million deaths each year [1]. Due to vigorous public health interventions, smoking prevalence in Australia has declined substantially over the last 50 years [2]. Nevertheless, 9.3% of the total disease burden (in disability-adjusted life years) was attributable to combustible tobacco use in 2015 [3].

E-cigarettes are a diverse group of battery-operated or rechargeable devices that heat a liquid (‘e-liquid’ or ‘e-juice’) to produce a vapour that users inhale. Although the composition of e-liquid varies, it typically contains a range of chemicals including propylene glycol and flavouring agents and are commonly used to deliver nicotine [4]. The labelling of electronic nicotine delivery systems (ENDS) and electronic non-nicotine delivery systems (ENNDS) is not always accurate, with reports of nicotine found in products labelled ENNDS [4, 5].

Studies indicate that in many countries, e-cigarette use among never-smoking youth is increasing [6-11]. In Australia, the proportion of non-smokers aged 14 years or older who had ever used e-cigarettes increased from 4.9% in 2016 to 6.9% in 2019 [12]. The increase was particularly notable in young adults, with 20% of 18-24 year old non-smokers reporting e-cigarette use [12].

There are concerns that the use of e-cigarettes in never-smokers may increase the probability that they will try combustible tobacco cigarettes and go on to become regular smokers, particularly among youth and young adults [13-15]. Furthermore, use of e-cigarettes could conceivably lead to combustible tobacco smoking relapse in former smokers. If e-cigarette use leads to more people smoking combustible cigarettes, compared with the number of people who have smoked in the absence of e-cigarettes, this would be a source of considerable public health harm [16]. Thus, our primary research question is: among current non-smokers, how does e-cigarette use affect the subsequent risk of smoking combustible tobacco cigarettes? This review aims to systematically update global contemporary population-level evidence on the relationship of e-cigarette use to smoking uptake.

## Methods

This summary of the global evidence comprises an umbrella review of systematic reviews and a top-up systematic review of primary research not included in the systematic reviews of the umbrella review. The protocol was published online through PROSPERO (CRD42020168596).

### Search strategy

The Population, Intervention, Comparison, Outcome (PICO) format was used to structure the search (Table in S1 Table). Studies investigating the association between ENDS or ENNDS use among non-tobacco smokers and uptake of combustible cigarette smoking were included. For both the umbrella review and the top-up systematic review, six databases (PubMed, Scopus, Web of Science, PsycINFO (Ovid), MEDLINE (Ovid), and Cochrane) were searched on 1 April 2020 (Text in S2). E-cigarette use, cigarette smoking and uptake related search terms and keywords were used (Table A in S2).

### Inclusion and exclusion criteria

Systematic reviews and meta-analyses of prospective cohort studies or randomised or non-randomised controlled trials examining the exposure (e-cigarette use) and outcome (smoking uptake in current non-smokers) of interest were included in the umbrella review. For the top-up systematic review, individual prospective cohort studies or randomised or non-randomised controlled trials identified in the search and not included in the umbrella review studies, were included. Cross-sectional studies were excluded due to difficulties in establishing the temporal relationship between e-cigarette exposure and smoking uptake. Studies with a follow up of less than 6 months or with abstracts not published in English were excluded. The full inclusion and exclusion criteria can be found in S2 Table B.

### Data screening and extraction

EndNote and Covidence software were used for review management. Two authors of this review (OB and LF) undertook initial screening, study selection, risk of bias assessment, and data extraction. Titles and abstracts identified in the searches were screened using a checklist, followed by full-text screening. A forward and backward reference search using Scopus was performed from the final included articles. After removing duplicates, titles, abstracts, and then full texts were screened for any studies fulfilling the inclusion and exclusion criteria. Data was independently extracted from the included systematic reviews and cohort studies using a pre-specified data extraction template. As it is important to consider whether authors of the studies under review hold any conflicts of interest that could potentially bias their findings, or whether the research was funded by an organisation with a financial interest in the outcomes, information on the source of research sponsorship or external involvement was also extracted. Studies were considered separately if they were funded by the tobacco or nicotine industry.

### Risk of bias assessment

Risk of bias for each study included was independently assessed using the AMSTAR 2[17] for the systematic reviews and meta-analyses included in the umbrella reviews, and the Newcastle Ottawa Scale (NOS)[18] for the studies in the top-up systematic review. For meta-analyses with at least ten studies, risk of bias across studies was assessed and interpreted using the symmetry of funnel plots and superimposed 95% confidence limits [19].

### Summary measures and synthesis of results

Findings from the umbrella review and the top-up systematic review were synthesised separately in narrative summaries. Individual prospective primary research studies identified from both the umbrella review and top-up systematic review were then considered in an integrated systematic review. Where appropriate, odds ratios from the studies in the integrated systematic review were combined using a random-effects model. Heterogeneity of study effect estimates were assessed by an I-squared statistic. All analyses were conducted using Stata version 16.1.

## Results

### Study Selection

Study selection for this umbrella review and top-up systematic review are shown in the PRISMA flowchart in Figure 1. A total of 6,225 studies were identified for title and abstract screening; 2,659 remained after exclusion of duplicates. After title and abstract screening, 83 articles were identified for full-text screening. Fifteen papers were identified for inclusion; three were systematic reviews that were included in the umbrella review and 12 were primary research studies included in the top-up systematic review. Ten of the latter studies were prospective observational studies and two were secondary analyses of randomised controlled trials (RCT).

**Fig 1:**
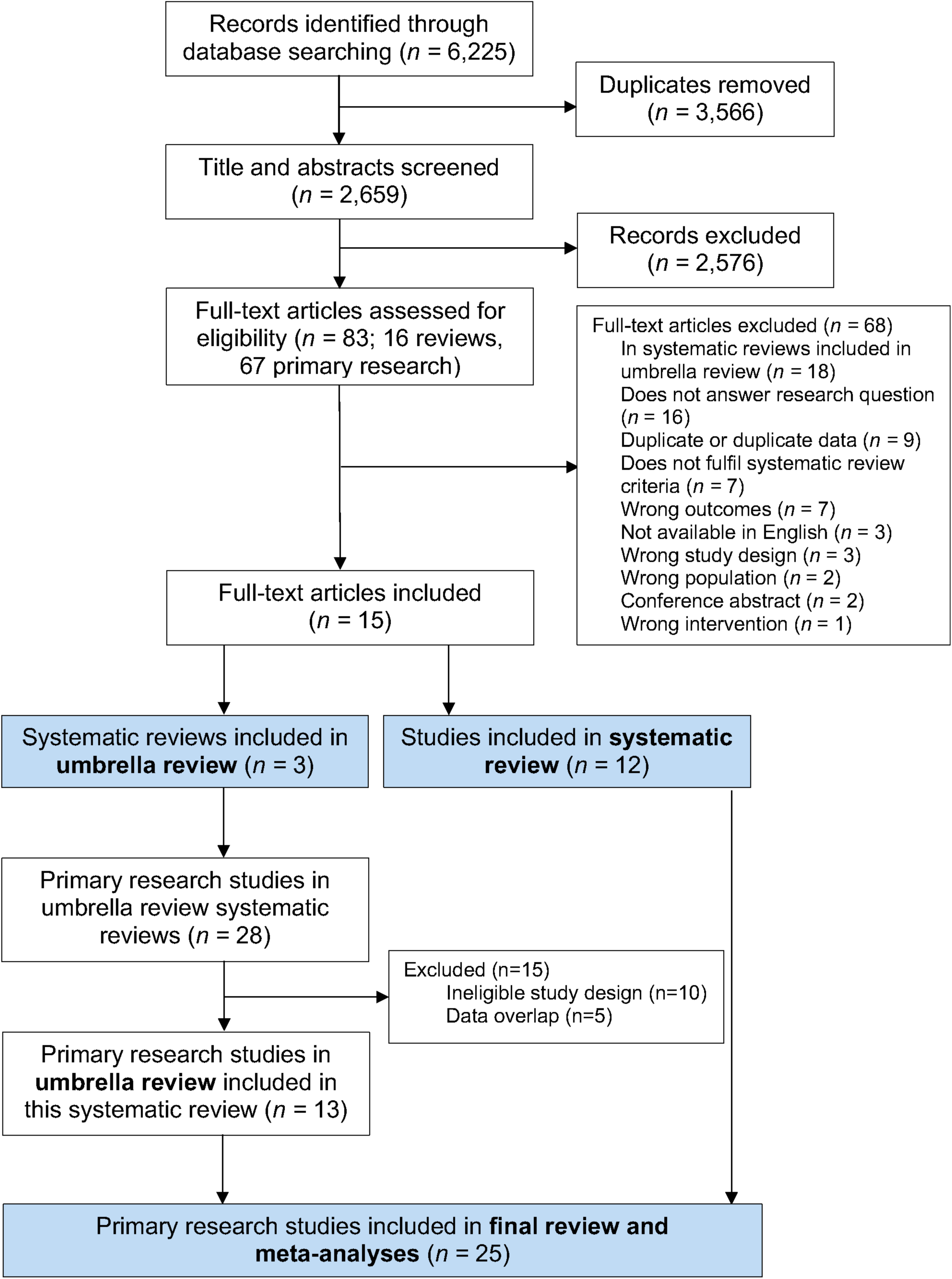
Flow chart for selection of studies for inclusion in umbrella review and top-up systematic review

From the three systematic review papers included in the umbrella review, 28 primary research studies were identified after removing duplicates. For our meta-analyses, we excluded 15 studies due to ineligible study design (n=10) or data overlap (n=5). No studies were excluded based on their quality assessment scores. The meta-analyses were thus based on 13 primary research studies identified from the prior systematic reviews, and 12 studies from our top-up systematic review, i.e. a total of 25 primary research studies on e-cigarette use and smoking uptake (Figure 1).

No potential competing interests were identified in the included studies themselves, or by the authors, based on the disclosure statements from the publications. Although one [20] primary research study identified during screening in the top-up systematic review was found to have potential competing interests, as it was funded by the tobacco industry, it was previously excluded due to a large overlap with data presented in a more recent paper by Berry et al. [21].

There is considerable uncertainty regarding the chemical constituents of the e-liquids delivered by the e-cigarettes in the studies included in the review. Where evidence on nicotine content was available, it indicated that a substantial majority of e-cigarettes in those studies delivered nicotine [22-25]. Many publications noted considerable uncertainty regarding nicotine content, including apparent mislabelling, and the need for greater clarity and reliability on this point.

### Umbrella review: quality assessment

All three systematic reviews from the selected articles rated moderate in the AMSTAR 2 [17] assessment. Information was lacking regarding study exclusion criteria, stated sources of funding, and detail on data extraction (Table in S3).

### Umbrella review

Table 1 summarises the results of the three systematic reviews included in the umbrella review. All three systematic reviews excluded studies with participants over 30 years of age. Sample sizes for the individual studies varied considerably, ranging from 298 to 17,318. Of the 13 included longitudinal primary research studies (detailed in Table in S4 Table), nine[14, 26-33] were based in the US, two [34, 35] in the UK and one each in Mexico [36], and the Netherlands [37]. Each of the three systematic reviews conducted meta-analyses and found the odds of smoking initiation were increased for youth and young adult e-cigarette users compared to non-e-cigarette users; these results are summarised in Table 1.

**Table 1:**
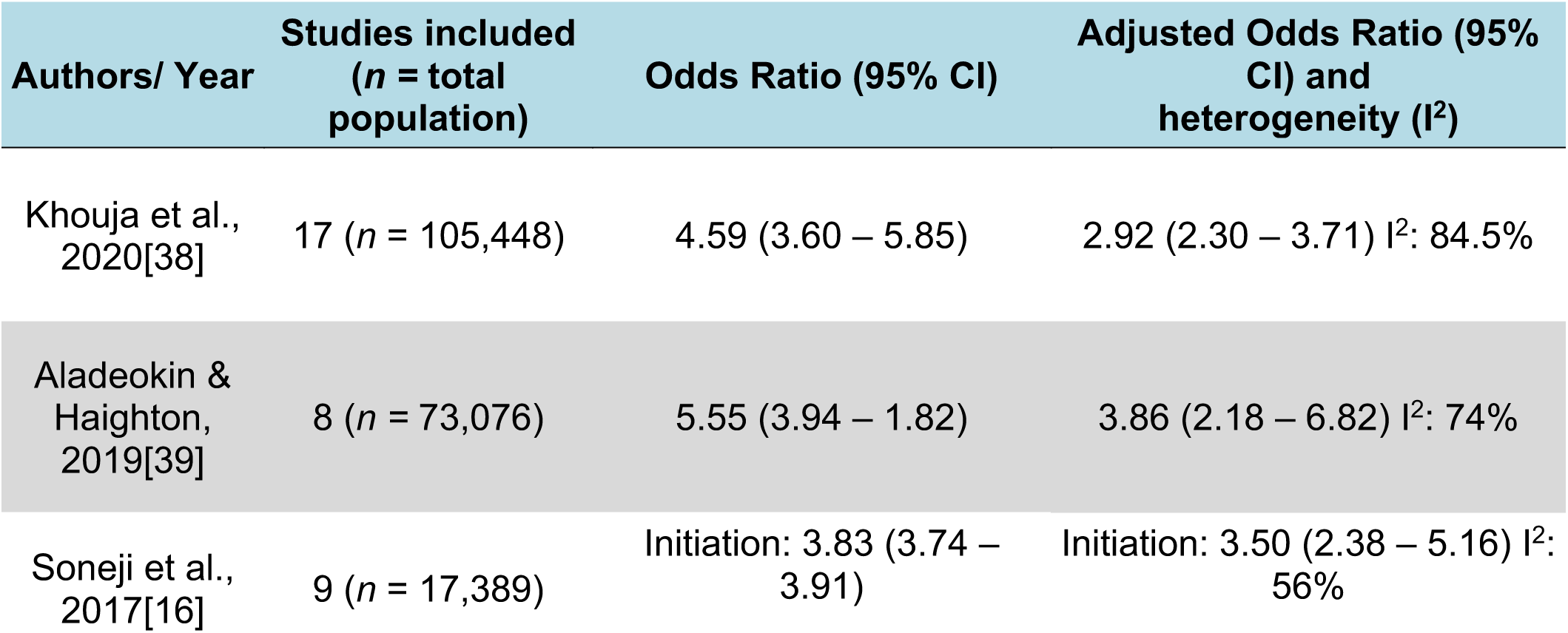
Odds ratios and adjusted odds ratios of the association between e-cigarette use and combustible cigarette smoking from systematic reviews and meta-analyses included in the umbrella review

The Khouja et al. (2020) systematic review and meta-analysis included 17 studies published up to November 2018 [38]. The study found that the risk of later smoking in people aged <30 years who had ever used or currently use e-cigarettes was strong; an almost three-fold the odds compared to never users after adjustment for covariates (see Table 1). However, there were high levels of heterogeneity in the summary estimates (adjusted OR I^2^ =84.5%), which remained high in adjusted analysis subgrouping by age, ever smoking, risk of bias and location of study. Heterogeneity was reduced when the adjusted ORs were grouped into those examining the relationship between ever e-cigarette use and current smoking (adjusted OR 2.21; 95% CI 1.72 – 2.84, I^2^ =5%) and those assessing the relationship of current e-cigarette use to ever smoking (adjusted OR 2.21; 95% CI 1.72 – 2.84, I^2^=5%).

Aladeokin & Haighton (2019) aimed to systematically review the evidence on e-cigarette use and initiation of cigarette smoking in adolescents (aged 10-19 years old) in the UK and included eight studies.[39] Their meta-analysis showed e-cigarette users were much more likely than non-users to go on to smoke combustible cigarettes, even after adjusting for covariates (see Table 1); the substantial heterogeneity in the summary estimate should be noted.

The Soneji et al. (2017) systematic review and meta-analysis included nine longitudinal studies of US participants ≤30 years of age [16]. Seven of the included studies assessed the association of baseline ever e-cigarette use with subsequent ever combustible cigarette use at follow-up among baseline never smokers. Soneji et al. also identified two studies that assessed baseline past 30-day e-cigarette use with subsequent past 30-day combustible cigarette use among those reporting no past 30-day use of cigarettes at baseline. The meta-analysis showed a markedly higher odds of combustible cigarette use in those who had used e-cigarettes (Table 1).

### Top-up systematic review: quality assessment

The quality of the included studies was evaluated using the Newcastle-Ottawa Scale [18] (NOS). Of the 12 studies, the NOS totals (out of 10 stars) ranged from 5 to 8 (Table in S5). Only one [40] study rated 5, five [23-25, 41, 42] rated 6, two [9, 43] rated 7 and four [21, 44-46] rated 8. No studies received a star for assessment of outcome. The main areas impacting the NOS scores were ascertainment of exposure and adequacy of follow up of cohorts (studies with less than 30% loss to follow up were considered adequate).

### Top-up systematic review and integration with primary research studies from the umbrella review

A total of 12 studies published in 2018, 2019 and 2020 were newly-identified for the top-up systematic review (Table 2; Table in S6). Among the 12 included, six were from the US, two from the UK, and one each from Romania, Finland, Taiwan and Canada. Study sample sizes varied considerably, ranging from 374 to 14,623.

**Table 2:** Odds ratios and adjusted odds ratios of the association between e-cigarette use and subsequent combustible cigarette use for: (1) never-smokers at baseline, (2) non-smokers^a^ (never or no current use) at baseline and (3) former smokers at baseline

| Authors/ Year | Country | Baseline cigarette use | E-cigarette use | Follow up cigarette use | Odds Ratio (95% CI) | Adjusted Odds Ratio (95% CI) |
| --- | --- | --- | --- | --- | --- | --- |
| **(1) Initiation in never smokers at baseline** |  |  |  |  |  |  |
| Berrv et al., 2019 [21] | US | Never | Ever | Ever |  | 4.09 (2.97 – 5.63) |
| Chien et al., 2019 [22] | Taiwan | Never | Ever | Ever | 2.44 (1.94 – 3.09) | 2.14 (1.66 – 2.75) |
| Conner et al., 2019 [42] | UK (England) | Never | Ever | Ever | 4.03 (3.33 – 4.88) | 2.78 (2.20 – 3.51) |
| McMillen et al., 2019 | US | Never | Current<sup>b</sup> | Ever | 16.4 (9.8 – 27.5) | 6.6 (3.7 – 11.8) |
| Pénzes et al., 2018 [41] | Romania | Never | Ever | Ever | 2.75 (1.52 – 4.96) | 3.57 (1.96 – 6.49) |
| **(2) Current use in non-smokers at baseline** |  |  |  |  |  |  |
| Aleyan et al., 2019 [23] | Canada | Non-smokers<sup>a</sup> | Current<sup>b</sup> | Current<sup>b</sup> |  | Wave 1 to 2: 1.54 (1.37 – 1.74)<br>Wave 2 to 3: 1.18 (1.08 – 1.29) |
| Barrington-Trimis et al., 2019 [43] | US | Never | Current<sup>b</sup> | Current<sup>b</sup> |  | NHW to dual use: 7.44 (3.63–15.3)<br>HW to dual use: 3.64 (1.62–8.18) |
| Bold et al., 2018 [40] | US | No current<sup>a</sup> | Current<sup>b</sup> | Current<sup>b</sup> |  | Wave 1 to 2: 7.08 (2.34 – 21.42)<br>Wave 2 to 3: 3.87 (1.86 – 8.06) |
| Conner et al., 2019 [42] | UK (England) | Never | Ever | Current<sup>b</sup> | 3.38 (2.72 – 4.21) | 2.17 (1.76 – 2.69) |
|  |  |  |  | Regular<sup>c</sup> | 3.60 (2.35 – 5.51) | 1.27 (1.17 – 1.39) |
| Kinnunen et al., 2019 [24] | Finland | Never | Ever nicotine-containing | Daily | 11.52 (4.91 – 27.01) | 8.50 (2.14–29.19)<br>With school clustering: 2.92 (1.09–7.85) |
|  |  |  | Ever non-nicotine containing |  | 1.88 (0.25 – 14.45) | 2.50 (0.25–12.05)<br>With school clustering: 0.94 (0.22 – 4.08) |
| McMillen et al., 2019 [45] | US | Never | Ever (not current) | Established<sup>d</sup> | 5.9 (1.7 – 20.7) | 2.5 (0.6 – 10.9) |
|  |  |  | Current<sup>b</sup> |  | 25.5 (10.6 – 61.4) | 8.0 (2.8 – 22.7) |
| Osibogun et al., 2020[44] | US | Non-smokers<sup>a</sup> | Current<sup>b</sup> | Regular<sup>c</sup> | Year 1: 16.4 (7.8 – 34.5)<br>Year 2: 11.1 (3.5 – 35.2) | Year 1: 5.0 (1.9 – 12.8)<br>Year 2: 3.4 (1.0 – 11.5) |
| **(3) Relapse in former smokers at baseline** |  |  |  |  |  |  |
| Brose et al., 2019 [25] | UK | ≥2-month ex-smokers | Ever | Ever | 1.52 (0.88 – 2.62) | 1.13 (0.61 – 2.07) |
|  |  |  | Non-daily |  | 3.32 (1.23 – 8.96) | 2.45 (0.85 – 7.08) |
| Dai et al., 2019 [46] | US | >12-month ex-smokers | Current<sup>b</sup> | Ever | 6.36 (4.49 – 9.00) | 2.00 (1.25 – 3.20) |
|  |  |  | Occasional |  | 5.79 (1.50 – 22.33) | 1.56 (0.34 – 7.14) |
|  |  |  | Prior |  | 9.68 (4.74 – 19.75) | 3.77 (1.48 – 9.65) |
| McMillen et al., 2019 [45] | US | ≥5years ex-smokers | Ever (not current<sup>b</sup>) | Ever | 5.4 (2.9 – 10.2) | 3.3 (1.6 – 6.7) |
|  |  |  | Current<sup>b</sup> |  | 7.6 (3.0 – 19.4) | 5.2 (1.6 – 16.3) |

NHW: Non-Hispanic White; HW: Hispanic White
a - non-smokers defined as never or no current (past 30-day) use
b - current defined as past 30-day use
c - regular defined as ≥20 days/ 30 days
d - established defined as ≥100 combustible cigarettes and currently smokes every day or some

Of the six newly-identified studies based on US participants, four[21, 44-46] used Population Assessment of Tobacco and Health (PATH) data from a US nationally representative longitudinal study. Of these, two [45, 46] looked at adult (≥ 18 years old) former smokers, one [44] looked at youth (12-17 years old) and one [21] at a more restricted youth group (12-15 years old). Even though these four studies have the same data source, they were all included in this review as they had different outcome or exposure variables, different populations and included the most recent data.

Of the 12 newly identified studies, five [21, 22, 41, 42, 45] had outcomes assessing ever smoking among never smokers at baseline, seven [23, 24, 40, 42-45] had outcomes assessing current smoking among non-smokers (never or not current smoking) at baseline and three [25, 45, 46] assessed the odds of relapse in former smokers. Results were separated based on these three categories and combined with the 13 primary research studies identified in the umbrella review. Twelve of the seventeen studies in Khouja et al. were included [14, 26, 28-37],three were excluded due to data overlap [47-49], one was excluded as it used retrospective data[50] and one was excluded as it was cross-sectional [51]. Of the eight studies in Aladeokin & Haighton, two were included [34, 35]; five were excluded for cross-sectional design [52-56] and one for data overlap [49]. From the nine studies identified in Soneji et al., six were included [26-29, 31, 32] after two were excluded as they were abstracts and one excluded for data overlap [57].

### Cigarette smoking initiation among never smokers at baseline

Five [21, 22, 41, 42, 45] of the newly-identified studies investigated smoking initiation among never smokers, of which Berry et al. [21] and McMillen et al. [45] used PATH data, focusing on youth (12 to 15 years old) and adults (≥18 years old), respectively (Table 2). Chien et al. examined the association between ever e-cigarette and subsequent combustible smoking initiation in 12,954 youth enrolled in schools in Taiwan between 2014 and 2016 [22]. Conner et al. investigated the association of e-cigarette use at baseline and smoking in adolescents (13 to 14 years old) between Waves 3 and 5 (2014 to 2016) of a cluster RCT in 20 schools in England [42]. Pénzes et al. conducted secondary data analysis from 1,369 9^th^ grade students in the Romanian ASPIRA randomized controlled trial. Details of the studies are given in S6.[41]

All newly-identified studies found that people who used e-cigarettes were significantly more likely than non-users to initiate smoking of combustible cigarettes, with odds ratios varying substantially from 2.1 to 6.6 (I^2^=81%; Figure 2a).

**Fig 2:**
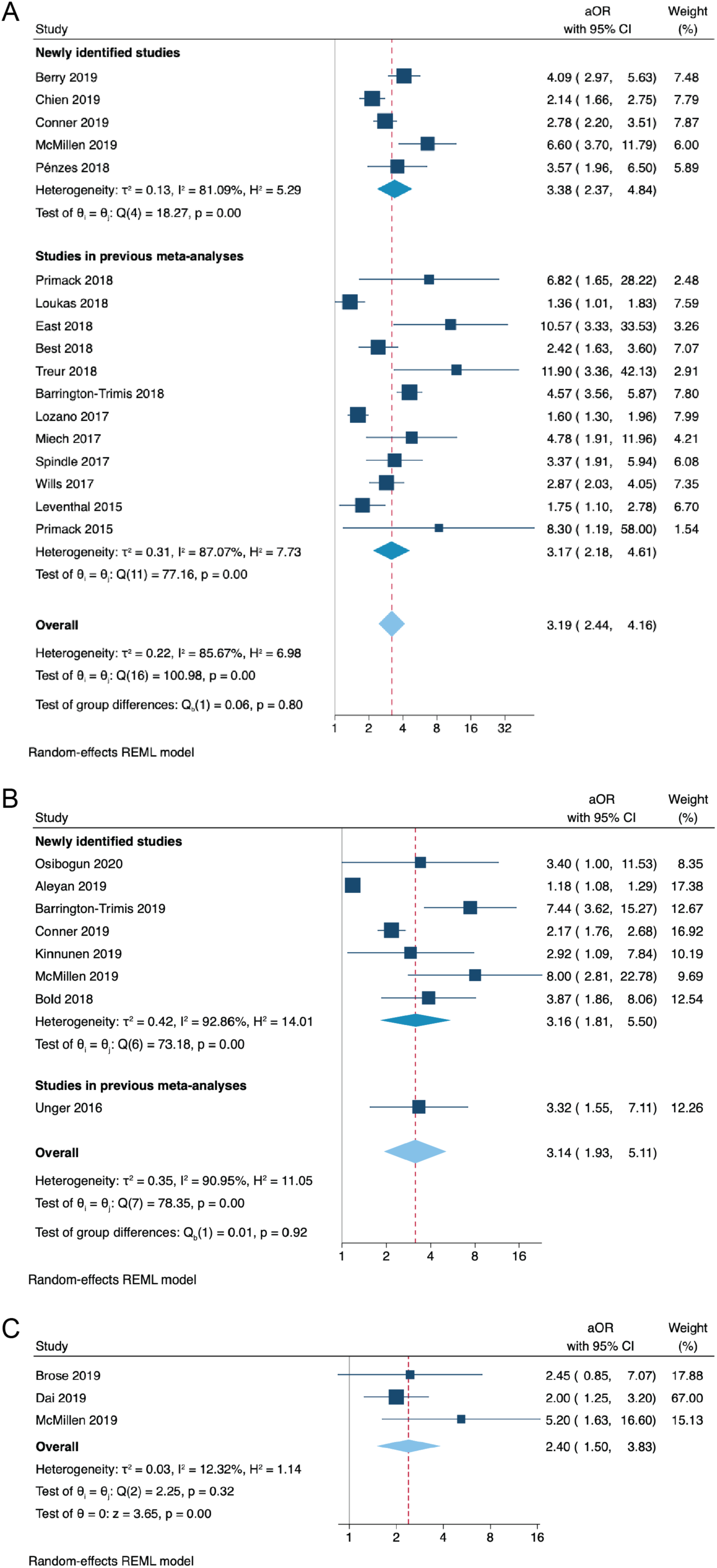
Forest plot and random-effects meta-analysis for the adjusted odds of; A) smoking initiation at follow up among never smokers and current e-cigarette users at baseline compared with never e-cigarette users at baseline B) current (past 30-day) smoking at follow up among non-current smokers and current e-cigarette users at baseline compared with non-current e-cigarette users at baseline C) smoking relapse at follow up among former smokers and current e-cigarette users at baseline compared with never e-cigarette users at baseline

Considering these newly identified studies along with 12 studies from the umbrella review, all found significantly increased risk of initiating smoking of combustible cigarettes in people who had used e-cigarettes, compared to those who had not (Figure 2a). Combining the studies from the umbrella review with the newly-identified studies, people exposed to e-cigarettes more likely to take up smoking of combustible cigarettes than people who were not exposed to e-cigarettes (pooled adjusted OR 3.19 (95% CI, 2.44 – 4.16)).

### Current (past 30-day) cigarette smoking among non-smokers (never smokers or no current use at baseline)

Seven [23, 24, 40, 42-45] of the newly-identified primary research studies investigated current (past 30-day) use of combustible cigarettes following the use of e-cigarettes (Table 2). Four [24, 42, 43, 45] of these studies looked at never smokers at baseline, while three [23, 40, 44] looked at non-smokers (either never or no current use).

Two [44, 45] of the included studies were based on PATH data. McMillen et al.[45] used data on adult (≥18 years old) never smokers from waves 1 to 2 of the PATH study and Osibogun et al. [44] used data on youth (12-17 years old) non-smokers from waves 1 to 3. A further two [40, 43] of the newly-identified studies used data from the US. Bold et al. surveyed 808 high school students across three waves (2013 to 2015) in Connecticut.[40] Barrington-Trimis et al. collated data on 6,258 youth from three US school-based studies between 2013 and 2015: the Children’s Health Study (CHS); the Happiness & Health Study (HH); and the Yale Adolescent Survey Study (YASS) [43]. This study separated results based on ethnicity and found the adjusted odds of dual use at follow up was considerably higher in non-Hispanic whites compared to Hispanic whites (see Table 2), although with considerable overlap in the confidence intervals

The remaining three [23, 24, 42] newly-identified studies used data from Canada, the UK and Finland. Aleyan et al. examined the association between current e-cigarette use and subsequent current smoking among 6,729 Canadian school students using data from a school-based longitudinal cohort study, COMPASS [23]. Conner et al. investigated the association of e-cigarette use at baseline and smoking between Waves 3 and 5 (2014 to 2016) of a cluster RCT assessing a self-regulation anti-smoking intervention from 20 schools in England [42]. Kinnunen et al. used MEtLoFIN a school-based longitudinal cohort dataset in 3,474 Finnish adolescents between 2014 and 2016 [24]. Kinnunen et al., separated the use of e-cigarettes using nicotine content and found among baseline never-smokers, ever use of nicotine-containing e-cigarettes was associated with a nearly 3-fold increase in the odds of uptake of daily smoking (see Table 2)) found no increase in risk associated with use of non-nicotine containing e-cigarettes.

All of the newly-identified studies, and the one relevant study from the umbrella review [27], found a significant increase in the risk of transitioning from being a non-smoker to a current smoker in people who had used e-cigarettes compared to not using e-cigarettes, but with considerable heterogeneity in the estimates (I^2^=91%; Figure 2b).

### Cigarette smoking relapse among former smokers (at least two months since quit date)

Three [25, 45, 46] newly-identified studies in this review investigated the odds of relapse to combustible cigarette smoking following the use of e-cigarettes in adults aged ≥ 18 years (Table 2). None of the three previously conducted systematic reviews investigated this relationship, so no additional studies from the umbrella review were included. Brose et al. used data from 371 adults who quit ≥2 months prior to baseline in 2016 from a national web-based survey in the UK [25]. The other two studies used PATH data. Dai et al. looked at 3,210 ex-smokers, who had not smoked for >12-months [46]. McMillen et al. looked at data relating to 8,108 adults who had quit ≥5 years prior to baseline; sub-analyses from this study were included in the previous two sections, as the study also provided data on never smokers [45].

All three included studies found the odds of ever relapse was higher among ever e-cigarette users, compared to never e-cigarette users (Figure 2c). Additionally, the odds of ever relapse was higher among current e-cigarette users than non-current e-cigarette users. A meta-analysis of the three newly identified studies found former smokers who used e-cigarettes had 2.4 times greater odds of relapse when compared to those who did not use e-cigarettes, with similar magnitudes of this relationship between studies (I^2^ = 12%) (Figure 2c).

### Risk of bias across studies

Funnel plots corresponding to the studies included in the meta-analyses are presented in Table in S7 Table. The plot for the seventeen smoking initiation studies of never-smokers is somewhat asymmetrical and seven points lie outside the 95% confidence region, suggesting there may be some selection bias across included studies, publication bias or possible heterogeneity (as supported by the I^2^ statistic; 86%). With less than ten studies investigating current smoking in non-smokers [23, 24, 27, 40, 42-44] and relapse in former smoke r[25, 45, 46], test for funnel plot asymmetry was not used as the power of the test would be too low for it to be a reliable indicator of publication bias [19].

## Discussion

Our umbrella and systematic review, along with an updated meta-analysis using data from primary studies, shows strong and consistent evidence that never smokers who have used e-cigarettes are more likely than those who have not used e-cigarettes to try smoking conventional cigarettes and to transition to become regular tobacco smokers. We found that, on average, non-smokers who used e-cigarettes have around three-fold the odds of either initiating smoking or currently smoking combustible cigarettes compared to non-smokers who have not used e-cigarettes. The limited available evidence indicates that former smokers who use e-cigarettes have more than twice the odds of relapse and resumption of current smoking compared to former smokers who have not used e-cigarettes.

This review builds on and has findings consistent with earlier systematic reviews and meta-analyses in the peer-reviewed and grey literature [11, 16, 38, 39, 58, 59]. A 2018 review by the National Academies of Sciences, Engineering, and Medicine (NASEM) on the public health consequences of e-cigarettes concludes that there is substantial evidence that e-cigarette use increases risk of ever using combustible tobacco cigarettes, and moderate evidence that e-cigarette use increases the frequency and intensity of subsequent combustible tobacco smoking, among youth and young adults [59]. Previous systematic reviews have focused on evidence in those 30 years of age or less, whereas our review included data on adults and former smokers. This is the first systematic review to examine whether e-cigarette use is associated with smoking relapse.

The use of e-cigarettes may represent a risk factor for cigarette smoking initiation, current smoking and relapse to cigarette smoking for several behavioural and physiological reasons. For those who use nicotine-containing e-cigarettes, a resulting addiction to nicotine may leave users at risk of seeking other forms of inhalable nicotine, such as combustible cigarettes [60, 61]. Additionally, as e-cigarettes can mimic behavioural (e.g. hand-mouth) and sensory (e.g. taste) aspects of smoking, associated e-cigarette habits and movements may make the transition to combustible smoking more natural [62, 63]. Further studies should examine potential mediators to better understand possible mechanisms for the association between e-cigarette use and subsequent cigarette use. Although one study showed that an intervention designed to reduce smoking initiation in adolescents through self-regulatory implementation intentions attenuated the odds of smoking uptake in never smokers who used e-cigarettes, a statistically significant increased odds remained [42].

Although studies in this review were consistent in finding increased risks of smoking uptake in non-smokers exposed to e-cigarettes, the magnitude of this increased risk varied substantially between studies. The reason for this variation is unclear, but may relate to the different products, populations and policy environments. In addition, it is challenging to estimate the overall effect of e-cigarettes on smoking initiation due to the variety of ways in which devices (e.g. e-cigarettes, JUULs, pods, vape pens) and users (e.g. never-users, ever-users, current-users, former users) are classified. The high heterogeneity in most of the results from the meta-analyses suggests that pooled odds ratios should be interpreted as an average of disparate results, rather than a reflection of the true underlying effect.

A limitation in this review is that included studies were limited to those written in English. While emerging results from this review and similar studies provide evidence regarding the association between e-cigarette and combustible cigarette use, the evidence is heavily weighted towards US and UK data. Only nine countries were included in this analysis, with a notable lack of data from the Asia-Pacific, Africa and the Middle East. Furthermore, the studies were reliant on self-reported product use, which is likely to be subject to self-reporting bias. All three systematic reviews rated moderate in the AMSTAR 2 risk of bias assessment and the 12 newly-identified studies rated between 5 to 8 on the Newcastle-Ottawa Scale. Although the consistency of findings across multiple studies and settings supports the likelihood of a causal relationship, given the observational nature of many of the included studies, the findings may be potentially influenced by confounding factors, including socioeconomic status and the tendency for risk behaviours to occur together. As the ability to adjust for such confounding factors varied according to study, the possibility of residual confounding cannot be excluded.

## Conclusion

This review found consistent evidence that use of e-cigarettes, largely nicotine-delivering, is associated with increased risk of subsequent combustible smoking initiation, current combustible smoking and smoking relapse after accounting for known demographic, psychosocial and behavioural risk factors. This is the first review to examine associations between e-cigarette use and cigarette use across the whole population, including youth, adults and former smokers. Intervention efforts and policies surrounding e-cigarettes are needed to reduce the potential of furthering combustible tobacco use in Australia.

## Data Availability

Supplementary material provided separately.

## Acknowledgments

RML is supported by a Senior Research Fellowship and EB is supported by a Principal Research Fellowship from the National Health and Medical Research Council of Australia.

